# Adverse events attributed to tracheal extubation in pediatric anesthesia: protocol for a multicenter prospective observational study (Ex-PEDIA; Extubation in Pediatric Anesthesia) in Japan

**DOI:** 10.64898/2026.01.06.26343502

**Authors:** Megumi Okuyama, Shogo Ichiyanagi, Tomoharu Ukiya, Katsuhiko Ishibashi, Fumio Watanabe, Shugo Kasuya, Yu Kaiho, Hiroshi Yonekura, Takahiro Imaizumi, Taiki Kojima

## Abstract

**Background:** Post-anesthetic tracheal extubation can lead to critical adverse events (AEs) in children due to their intolerance to hypoxia. However, evidence from pediatric studies on the factors associated with safe tracheal extubation is limited. We will investigate the incidence and risk factors of AEs associated with tracheal extubation using real-world data from Japan. This study aims to test the following hypotheses: extubation-related AEs are associated with (a) several indicative signs of cranial nerve activity and upper airway reflexes, and (b) difficult airway features.

**Methods:** This prospective, multi-center, registry-based, cross-sectional study will be conducted at 17 hospitals in Japan from October 2025 to September 2028. It will include children aged <18 years undergoing surgical and/or diagnostic test procedures under general anesthesia or sedation by anesthesiologists. Data on the patient, surgery, and anaesthesia characteristics; provider discipline; airway management; and extubation methodology will be collected. The exposures of interest are the presence of clinical signs related to anesthesia emergence and features of difficult airway. The primary and secondary endpoints are AEs associated with extubation and reduced peripheral arterial oxygen saturation. Multilevel mixed-effects multivariable regression analysis will be performed to adjust for potential confounders associated with AEs attributed to extubation and variations related to the hospital type and institutional level. The required sample size was determined to be approximately 8500 based on the assumptions of a 99% probability of obtaining a 95% Wilson CI with a half-width of ±0.3% and an incidence of any AE of 2.0%.

**Discussion:** This study is a prospective, registry-based, multicenter, cross-sectional study designed to describe the real-world incidence of AEs related to extubation and the associated risk factors in Japan. This study employs the validation system incorporating site-specific leaders and the REDCap® data registration system to minimize reporting and selection bias. We have conducted a pilot study to investigate feasibility and consulted research members regarding data collection and study methods. The findings will provide vital information on the risks associated with extubation-related AEs and contribute to the development of safer tracheal extubation strategies.

**Trail registration number:** jRCT 1030250100

## Background

Airway management under anesthesia contributes to life-threatening adverse events (AEs) in children, especially during the awakening period [1,2]. The mechanisms underlying the processes of emergence from anesthesia have not been fully elucidated, and this is attributable to the complex mechanisms underlying the recovery of consciousness and airway reflexes [3,4]. Strategies for determining the appropriate timing of tracheal extubation (TE) have also not been standardized. Therefore, the timing of extubation varies for anesthesia providers.

Pediatric airway management research databases have been established in Europe and Western countries [2,5]. However, reports on the strategies for post-anesthetic TE in the Asian region are limited. An observational study reported several awakening signs predictive of extubation success, but the findings were limited by the single-center design, focus on inhalational anesthesia, and restriction to awake TE [6]. The presence of difficult airway features has been associated with increased extubation failure and reintubation [7]. However, reports in the literature on the association between difficult airway features and respiratory AEs are still limited.

We will explore the incidence of AEs resulting from TE during anesthesia emergence in the Japanese pediatric population. This study aims to test two hypotheses about the predictors of extubation-related AEs: (a) AEs are associated with several indicative signs of cranial nerve activity and recovery of upper airway reflexes; and (b) AEs are associated with the presence of difficult airway features.

## Methods

### Study design and setting

Data for the prospective, registry-based, multi-center, real-world, cross-sectional study will be collected from October 2025 to September 2028 from 17 tertiary care hospitals in Japan. The centers include 6 pediatric hospitals, 7 mixed pediatric–adult university hospitals, and 4 mixed pediatric-adult community hospitals. These centers are categorized as follows:

Pediatric hospitals: National Center for Child Health and Development, Aichi Children’s Health and Medical Center, Hyogo Prefectural Children’s Hospital, Osaka Women’s and Children’s Hospital, Okinawa Prefectural Southern Medical Center & Children’s Medical Center, and Saitama Children’s Medical Center.

Mixed pediatric–adult university hospitals: Institute of Science Tokyo, Kitasato University Hospital, Nara Medical University, Tokyo Women’s Medical University, Yamagata University Hospital, Kochi Medical School Hospital, and Tohoku University Hospital.

Mixed pediatric–adult community hospitals: Matsudo City General Medical Center, Teina Keijinkai Hospital, Kimitsu Chuo Hospital, and Chiba City Kaihin Hospital.”

## Target population

TE is defined as the removal of the tracheal tube during emergence after surgery, when the patients no longer require any airway management.

## Inclusion criteria

The inclusion criteria were as follows: (a) children aged <18 years undergoing general anesthesia or sedation for scheduled or emergency surgery and/or test procedures conducted by anesthesiologists or anesthesia providers under supervision; (b) patients undergoing airway management with tracheal intubation (TI) and having the tracheal tube removed after the surgery and/or test; and (c) patients undergoing TE performed in operating suites, catheterization laboratory rooms, rooms for radiation imaging and procedures (such as CT, MRI, radiation therapy), or the general ward.

The following patients will be excluded: (a) children who will not receive TI; (b) patients who have undergone tracheostomy or those scheduled to undergo tracheostomy; (c) patients scheduled to continue mechanical ventilation under sedation after surgery and/or test procedures; (d) patients for who several extubation attempts have been made by the same personnel during the study period; and (e) patients who (or those whose families) will refuse to provide relevant medical information for this study.

## Data collection

The registry-based cross-sectional study will prospectively collect data on the characteristics of the patients and surgery, disciplines of the anesthesia providers who decide on tracheal tube removal, airway management during anesthesia induction, patient condition before emergence from anesthesia, interventions before and after extubation, occurrence of AEs, and treatments for AEs (Tables 1–3).

**Table 1.**
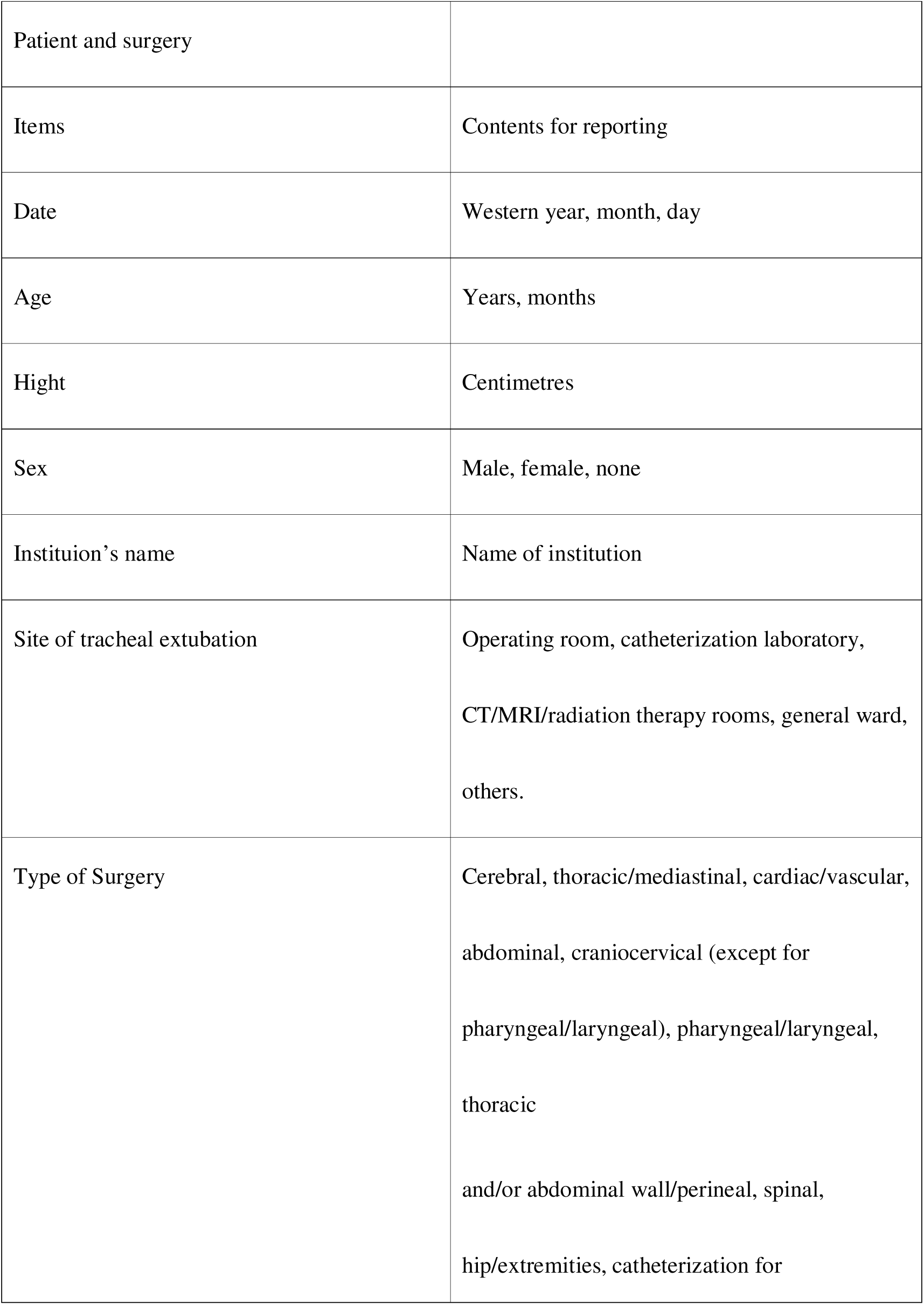
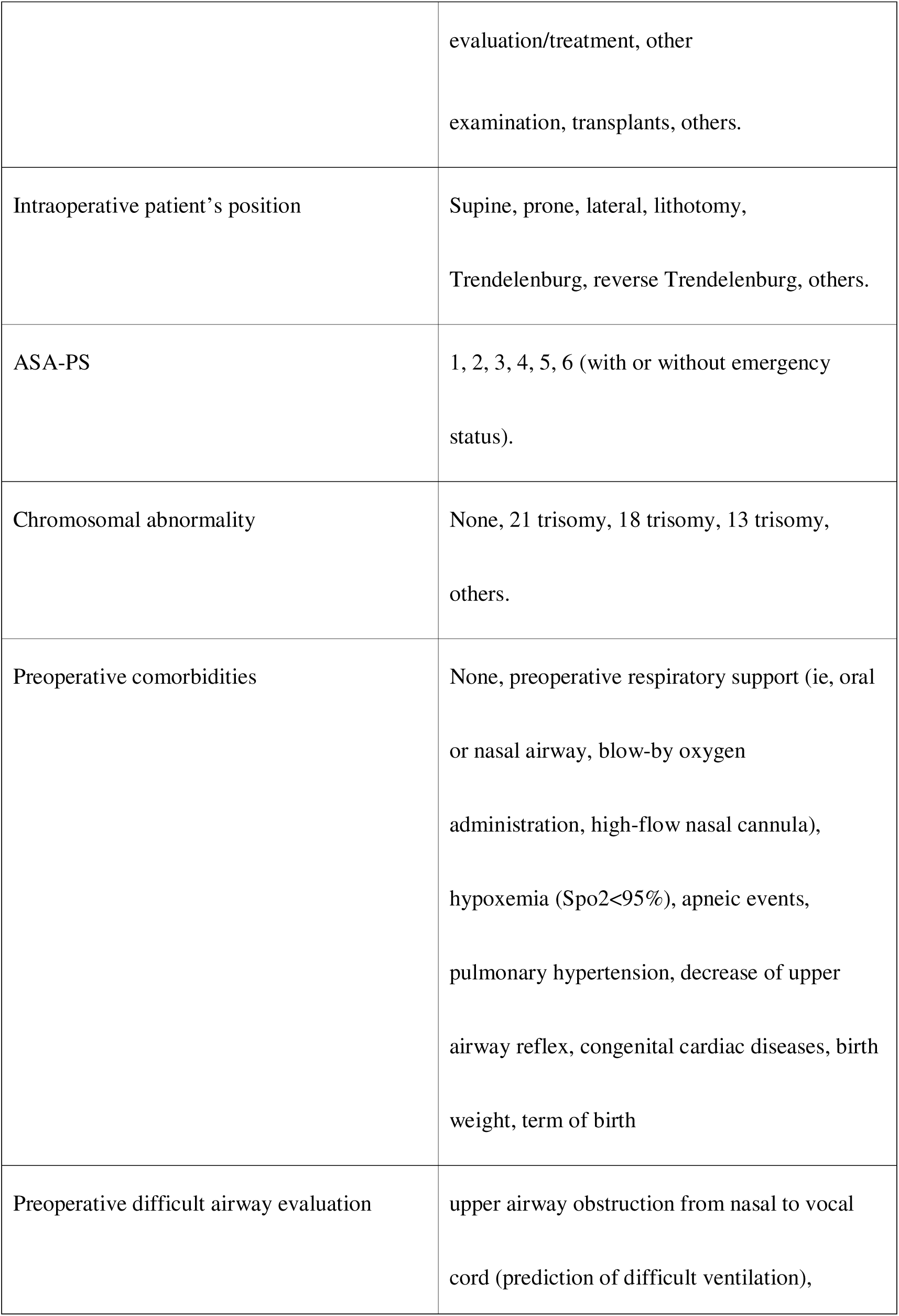
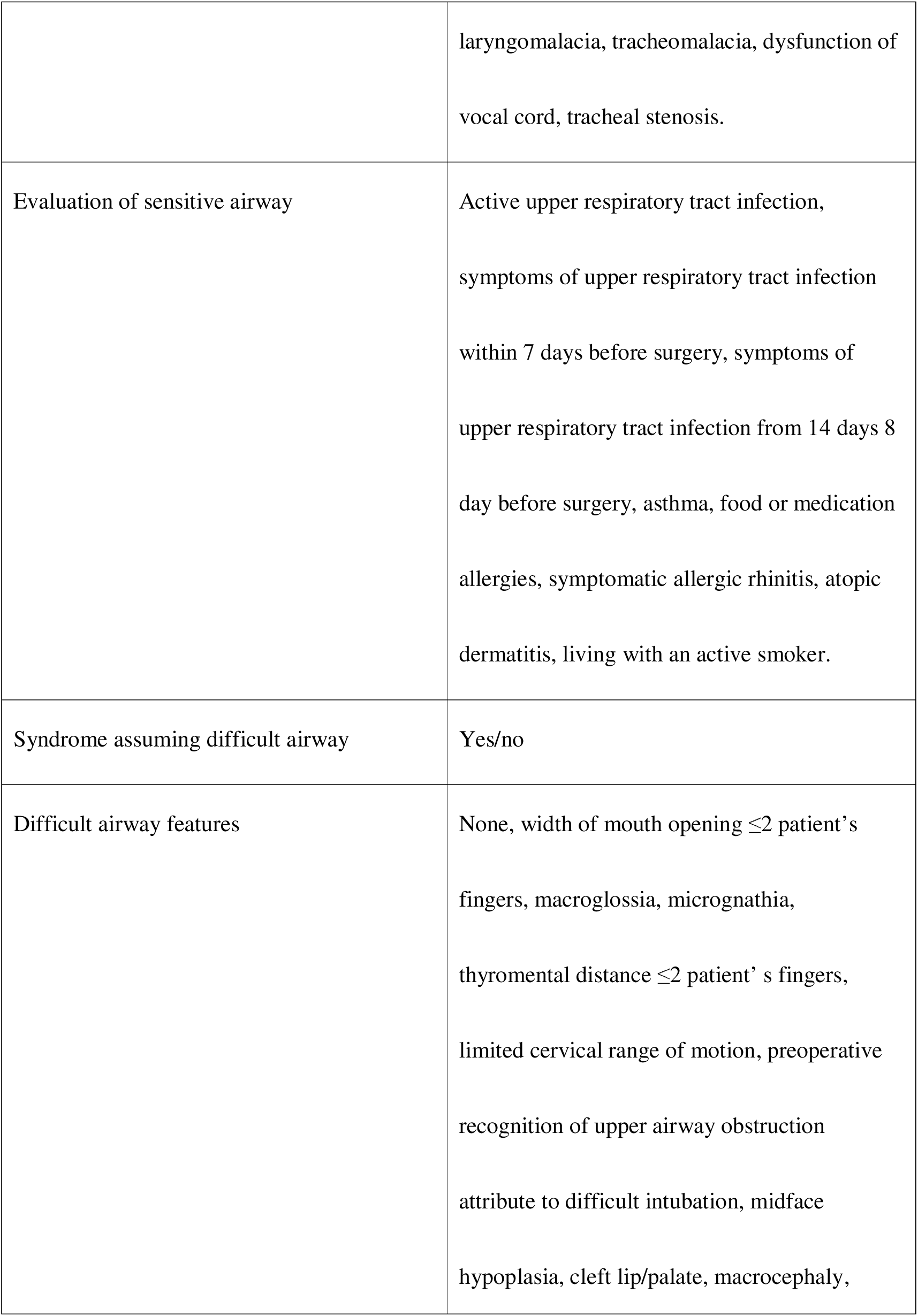
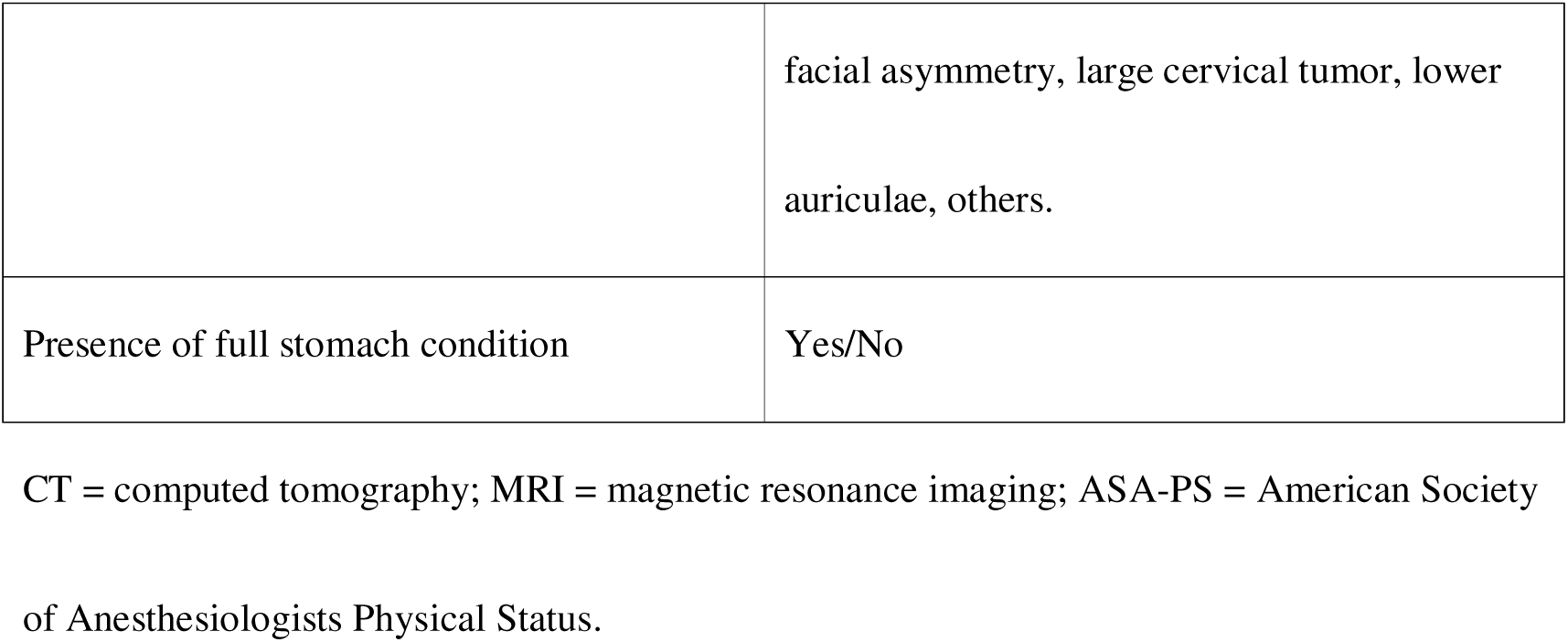
Details of the data collection forms regarding characteristics of the patients and surgery.

|  |  |
| --- | --- |
| Patient and surgery |  |
| Items | Contents for reporting |
| Date | Western year, month, day |
| Age | Years, months |
| Hight | Centimetres |
| Sex | Male, female, none |
| Instituion's name | Name of institution |
| Site of tracheal extubation | Operating room, catheterization laboratory,<br>CT/MRI/radiation therapy rooms, general ward,<br>others. |
| Type of Surgery | Cerebral, thoracic/mediastinal, cardiac/vascular,<br>abdominal, craniocervical (except for<br>pharyngeal/laryngeal), pharyngeal/laryngeal,<br>thoracic<br>and/or abdominal wall/perineal, spinal,<br>hip/extremities, catheterization for |
|  | evaluation/treatment, other<br>examination, transplants, others. |
| Intraoperative patient's position | Supine, prone, lateral, lithotomy,<br>Trendelenburg, reverse Trendelenburg, others. |
| ASA-PS | 1, 2, 3, 4, 5, 6 (with or without emergency status). |
| Chromosomal abnormality | None, 21 trisomy, 18 trisomy, 13 trisomy, others. |
| Preoperative comorbidities | None, preoperative respiratory support (ie, oral or nasal airway, blow-by oxygen administration, high-flow nasal cannula), hypoxemia (Spo2<95%), apneic events, pulmonary hypertension, decrease of upper airway reflex, congenital cardiac diseases, birth weight, term of birth |
| Preoperative difficult airway evaluation | upper airway obstruction from nasal to vocal cord (prediction of difficult ventilation), |
|  | laryngomalacia, tracheomalacia, dysfunction of vocal cord, tracheal stenosis. |
| Evaluation of sensitive airway | Active upper respiratory tract infection, symptoms of upper respiratory tract infection within 7 days before surgery, symptoms of upper respiratory tract infection from 14 days 8 day before surgery, asthma, food or medication allergies, symptomatic allergic rhinitis, atopic dermatitis, living with an active smoker. |
| Syndrome assuming difficult airway | Yes/no |
| Difficult airway features | None, width of mouth opening $\leq 2$ patient's fingers, macroglossia, micrognathia, thyromental distance $\leq 2$ patient's fingers, limited cervical range of motion, preoperative recognition of upper airway obstruction attribute to difficult intubation, midface hypoplasia, cleft lip/palate, macrocephaly, |
|  | facial asymmetry, large cervical tumor, lower auriculae, others. |
| Presence of full stomach condition | Yes/No |

CT = computed tomography; MRI = magnetic resonance imaging; ASA-PS = American Society
of Anesthesiologists Physical Status.

**Table 2.**
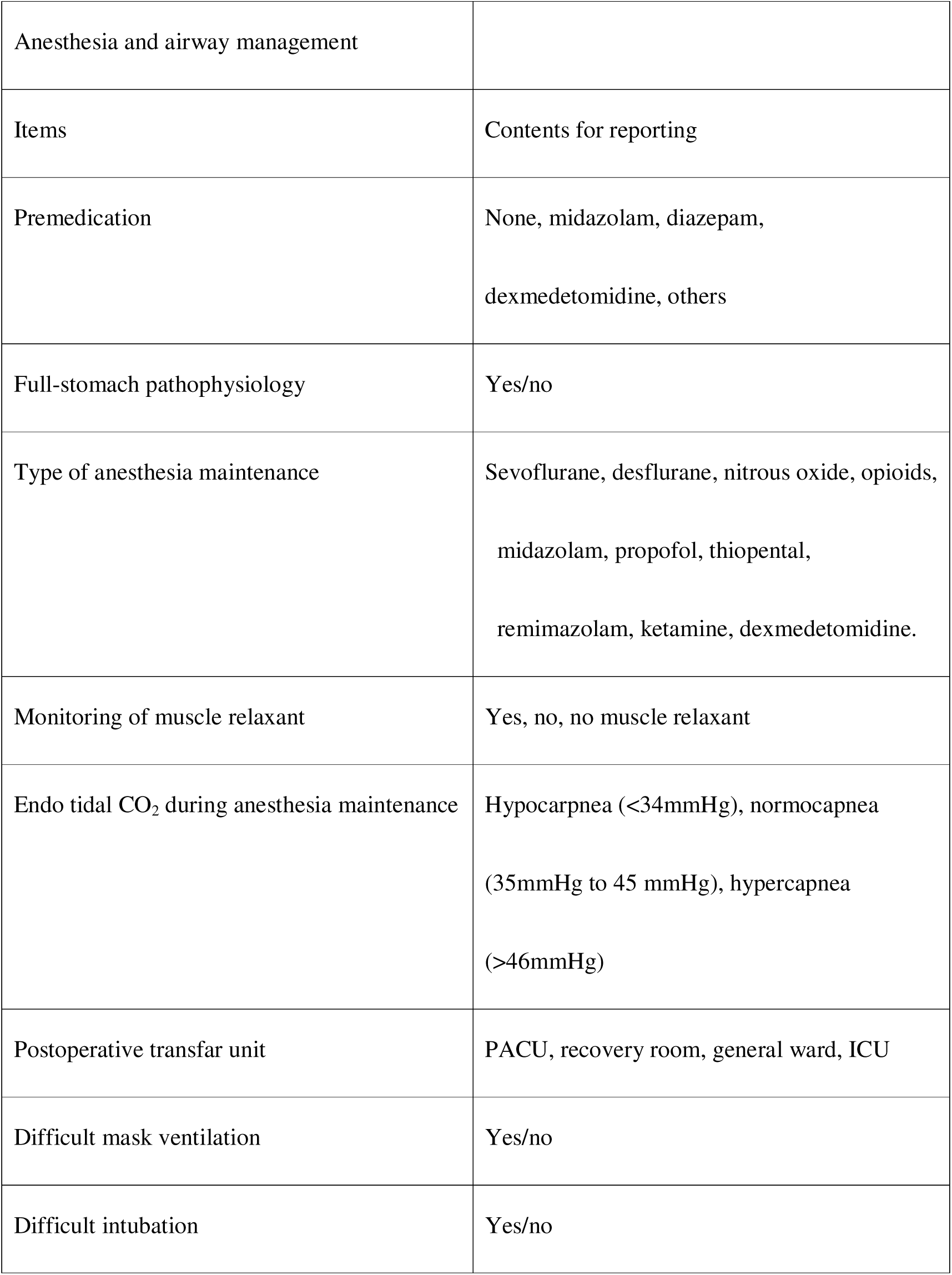
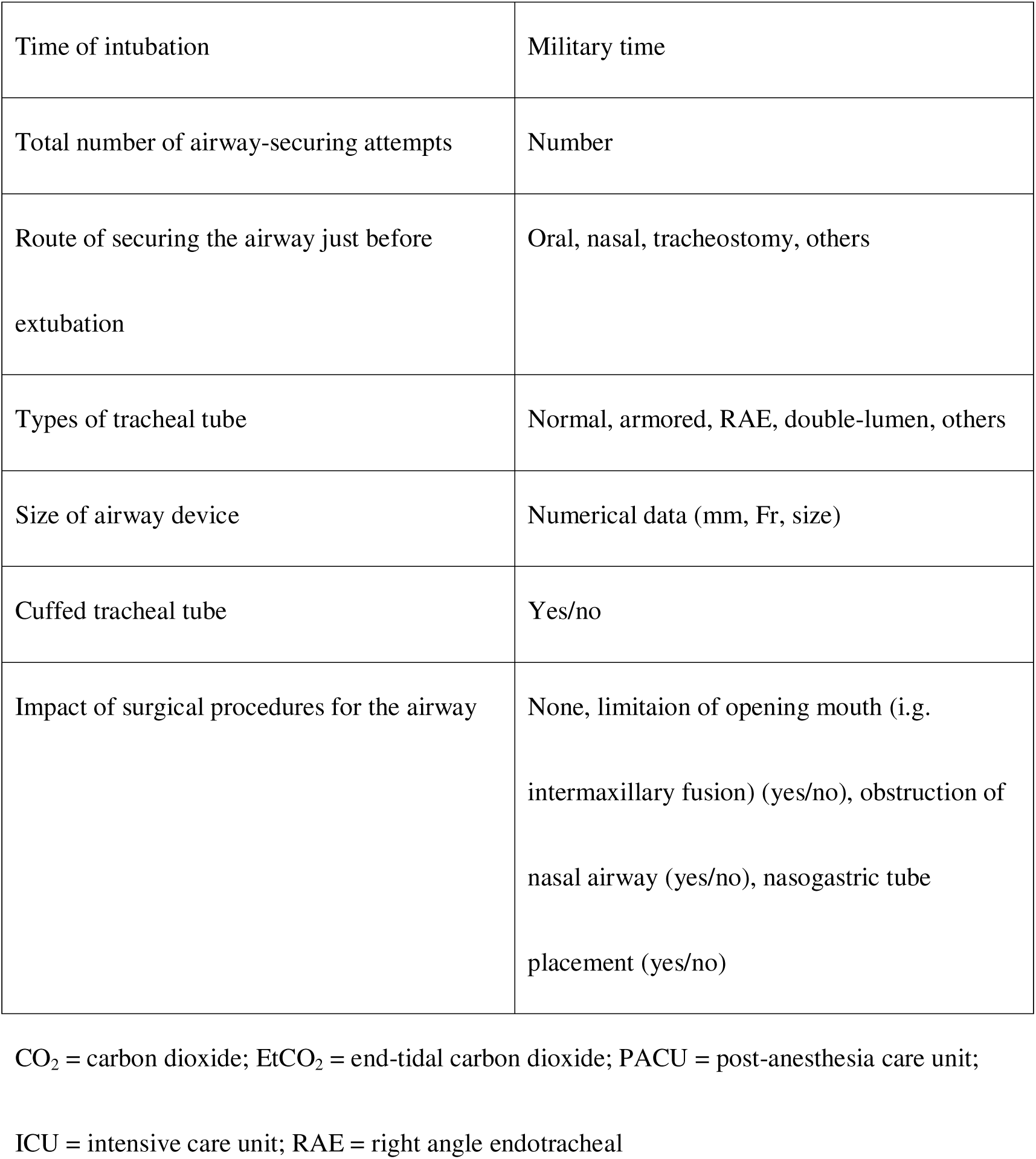
Details of the data collection forms regarding anesthesia and airway management.

**Table 3.** Details of the data collection forms regarding tracheal extubation.

|  |  |
| --- | --- |
| Extubation |  |
| Items | Contents for reporting |
| Briefing before extubation | None, among physicians, among physicians and nurses. |
| Use of muscle relaxant monitoring | Yes, no, not applicable (no use of muscle relaxants). |
| Administration of reversal medications | None, sugammadex, flumazenil, naloxone, vagostigmine/atropine. |
| Pretreatment for extubation | None, ☐-stimulant inhalant, airway placement, shoulder role placement, exchange airway equipment (e.g., tracheal tube to supraglottic airway device). |
| Oxygen administration during extubation | None, nasal cannula (amount of oxygen L/minute), high-flow nasal cannula (amount of oxygen L/minute), oxygen administration via nasal airway (amount of oxygen L/minute). |
| Decider the timing of extubation | Residents (junior or senior), fellows (anesthesia or other), board-certified anesthesiologists, other board-certified physicians, nurse anesthetists, board-certified dental anesthetists, others. |
| Presence of board-certified anesthesiologists on extubation | Yes/no. |
| Suction of secretion before extubation | None, oral cavity, nasal cavity, trachea. |
| Clinical sign predicting possible extubation | None, regular rhythm spontaneous breathing, tidal volume $\geq 5$ mL/kg, absence of spontaneous breathing, eye opening, grimacing, purposeful movement, tearing, movement without coughing, swallowing, coughing or cough-like reflexes. |
| Check for eye position before extubation | Unconfirmed, center, deviation. |
| Procedure of Laryngeal Stimulation Test | Unconfirmed, gentle cranial/caudal direction movement of endotracheal tube, suction of |
|  | pharynx, stimulation by deflation of the tracheal tube cuff, othres. |
| Reaction for laryngeal stimulation test | Spontaneous breathing maintained without disruption in capnometry, disruption in capnometry which immediately return to baseline wave forms within 5 seconds, disruption in capnometry which continues $\geq 5$ seconds, apnea continue $\geq 5$ seconds (flat in capnometry). |
| Any leak around tracheal tube after deflation | None, existence, unconfirmed. |
| Methods of extubation | Under positive airway pressure, under atmospheric pressure, under negative airway pressure (i.e., endotracheal suctioning). |
| Endtidal CO<sub>2</sub> before extubation | MmHg |
| Respiratory rate before extubation | Beat per minutes |
| Endtidal SEV/DES concentration before extubation | % |
| Adverse events between the completion of surgery and before extubation | None, cough, asthma attack, unplanned extubation, others. |
| Adverse events between the moment of extubation and leaving the operation room | None, cough lasting <10 seconds without apnea lasting for $\geq 10$ seconds, cough lasting <10 seconds with apnea lasting for $\geq 10$ seconds, cough lasting $\geq 10$ seconds, upper airway obstruction, laryngospasm, asthma attack, stridor, apnea lasting $\geq 10$ seconds, atelectasis/negative pressure pulmonary edema, vomiting, airway trauma, accidental extubation, pneumothorax/ pneumomediastinum, cardiac arrest/ severe bradycardia, arrhythmia. |
| Interventions for adverse events | None, jaw thrust, CPAP, positive pressure ventilation, shoulder roll placement, reposition of the patient (lateral, prone), oral suctioning, tracheal suctioning, airway (oral, nasal) insertion, reintubation, high-flow nasal canula |
|  | use, chest compression, ECMO attachment, defibrillation/ cardioversion, unplanned ICU admission. |
| Medications administered for adverse events | None, propofol, other sedatives, opioids, rocuronium/suxamethonium, inhalational bronchodilator, intravenous steroid, inhalational epinephrine, intravenous epinephrine, atropine, other vasopressors, anti-arrhythmic medications, sugammadex, flumazenil, naroxone, neostigmine/atropine. |
| SpO<sub>2</sub> value obtained at the completion of surgery | % |
| Lowest SpO<sub>2</sub> value obtained between the completion of surgery and immediately before extubation | % |
| Lowest SpO<sub>2</sub> value obtained between the initiation of extubation and when the | % |
| patients leaves the operating room |  |
| Type of extubation planned before<br>emergence | Awake extubation, deep extubation, unawake<br>extubation with spontaneuos breathing. |
| Type of extubation actual performed | Awake extubation, deep extubation, unawake<br>extubation with spontaneous breathing. |

CPAP = continuous positive airway pressure; ECMO = extracorporeal membrane oxygenation;
ICU = intensive care unit.

## Endpoints

The primary endpoint is the occurrence of an extubation-related AEs (dichotomous variable). The secondary endpoints are the presence of respiratory AEs and desaturation (decrease in SpO_2_ by ≥10%) (dichotomous variables).

The definitions of AEs and desaturation in the J-PEDIA study apply to this study [8]. The criteria for extubation-related AEs include at least one of the following conditions: severe cough lasting ≥10 seconds, upper airway obstruction, laryngospasm, bronchospasm, stridor, respiratory suppression (pause of respiration ≥10 seconds), atelectasis, negative pressure pulmonary edema diagnosed with imaging tests, vomiting, airway trauma, accidental extubation, pneumothorax, pneumomediastinum, cardiac arrest, and arrythmia including bradycardia. Respiratory AEs include at least one of the following conditions: severe cough lasting ≥10 seconds, upper airway obstruction, laryngospasm, pneumothorax, pneumomediastinum, bronchospasm, atelectasis, pulmonary edema, stridor, and airway trauma. We defined laryngospasm as complete airway obstruction associated with muscle rigidity of the abdominal and chest walls, and defined bronchospasm as increased respiratory effort, especially during expiration, and wheeze on auscultation (Table 3, Figure 1).

**Figure 1:**
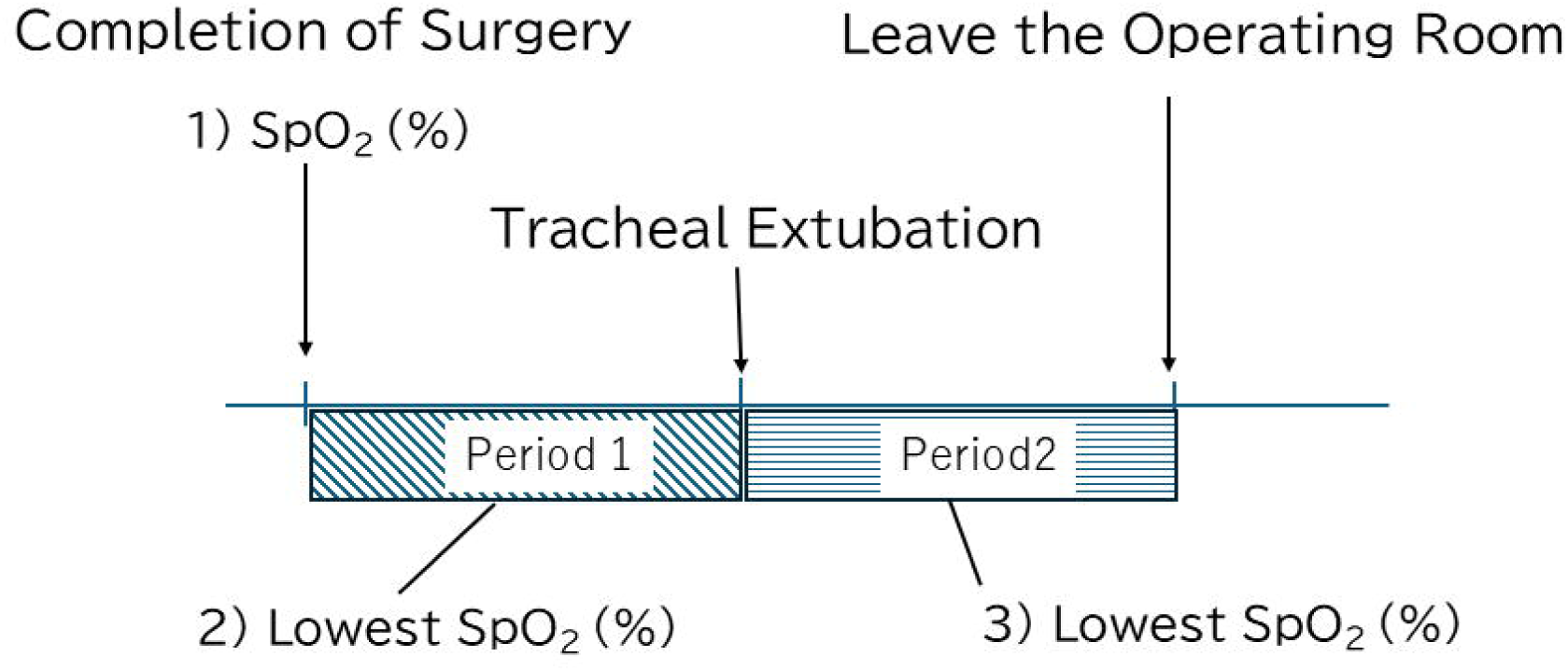

Three SpO_2_ values will be recorded by the anesthesia providers assigned to the cases: (a) value obtained at the completion of surgery; (b) lowest value obtained between the completion of surgery and immediately before extubation; and (c) the lowest value obtained between the initiation of extubation and when the patient leaves the operating room (Table 3, Figure 1). Desaturation is defined as a decrease in SpO_2_ of ≥10% after the surgery from the baseline value. It will be assessed during the period between the completion of surgery and immediately before extubation and that between the initiation of extubation and when the patient leaves the operating room.

## Exposures and potential confounders

The exposures are as follows: (a) presence of any clinical findings of activated central nerve functions, including spontaneous breathing with a regular rhythm, tidal volume >5 mL/kg, eye opening, grimacing, purposeful movement, tearing, movement without coughing, swallowing, coughing or cough-like reflexes, and conjugate gaze; (b) presence of any perioperative difficult airway features, including width of mouth opening of ≤2 fingers, micrognathia, macroglossia, thyromental distance of ≤2 fingers, limited cervical range of motion, upper airway obstruction, anatomical abnormality causing difficult laryngoscopy (such as tumor in mouth, soft tissue hypertrophy), midface hypoplasia, untreated cleft lip and/or palate, macrocephaly, morphologic asymmetries of the face, cervical tumor, lower auriculae, difficult airway syndromes, difficult mask ventilation at anesthesia induction, difficult intubation at anesthesia induction, limitation of mouth opening after surgery, obstruction of the nasal airway after surgery, and multiple attempts to secure the airway.

The potential confounders listed below were selected based on a review of the literature and the clinical knowledge of the anesthesiologists on the research team [2,5,6,9]. They will be adjusted to evaluate the following hypotheses: (a) association between the occurrence of AEs and clinical signs of activated central nervous systems, and (b) association between the occurrence of AEs and preoperative difficult airway features (Tables 4 and 5).

**Table 4.** Confounders for regression analysis.

| Association between post-extubation AEs and several indicative signs of cranial nerve activity and recovery of upper airway reflexes |  |
| --- | --- |
| Patient background | Sex (F/M), age (years), body weight (kg), ASA-PS (1,2,3,4,5), the presence of full stomach pathology (Y/N), birth abnormality<sup>1</sup> (Y/N), airway sensitivity<sup>2</sup> (Y/N), environmental sensitivity<sup>3</sup> (Y/N), type of surgery, laryngeal or pharyngeal dysfunction (Y/N), the presence of any difficult airway features<sup>4</sup> (Y/N), emergency surgery (Y/N) |
| Anesthetic management | Type of anesthetics for anesthesia maintenance (TIVA/GA), reversal of muscle relaxants (Y/N), adverse reversal of anesthetic (Y/N), anesthesia provider's discipline, endtidal CO<sub>2</sub> before extubation (mmHg) |
| Airway management | Duration of tracheal intubation (minutes), total number of attempts to secure the airway (times), oxygenation during extubation (Y/N), and oral or tracheal suction before |
|  | extubation (Y/N) |

<sup>1</sup> Birth abnormality, including preterm birth and low birth weight.
<sup>2</sup> Airway sensitivity, including active or within 14 days upper respiratory infection (URI)
symptoms, asthma, living with an active smoker.
<sup>3</sup> Environmental sensitivity, including food or medication allergies, allergic rhinitis, atopic
dermatitis.
<sup>4</sup> The presence of any difficult airway features including width of mouth opening of $\leq 2$ patient's
fingers, micrognathia, macroglossia, thyromental distance of $\leq 2$ patient's fingers, limited
cervical range of motion, upper airway obstruction, anatomical abnormality causing difficult
laryngoscopy, midface hypoplasia, untreated cleft lip and/or palate, macrocephaly, morphologic
asymmetries of the face, cervical tumor, lower auriculae, any difficult airway syndromes,
difficult mask ventilation or intubation at anesthesia-induction, limitation of mouth opening

**Table 5.** Confounders for regression analysis.

| Association between post-extubation AEs and the presence of difficult airway features |  |
| --- | --- |
| Patient background | Sex (F/M), age (years), body weight (kg), ASA-PS (1,2,3,4,5), the presence of full stomach pathology (Y/N), birth abnormality<sup>1</sup> (Y/N), airway sensitivity<sup>2</sup> (Y/N), environmental sensitivity<sup>3</sup>, type of surgery, place to perform extubation, type of surgery, position (supine, prone, others), emergency surgery (Y/N) |
| Anesthetic management | Premedication (none, midazolam, diazepam, others), type of anesthetics for anesthesia maintenance (TIVA/GA), muscle relaxant monitoring use (Y/N), reversal of muscle relaxants (Y/N), anesthesia provider's discipline, endtidal CO<sub>2</sub> before extubation (mmHg) |
| Airway management | Duration of tracheal intubation (minutes), advanced exchange of tube (Y/N), oxygenation during extubation (Y/N), total number of attempts to secure the airway (times), indicative signs of cranial nerve activity and |
|  | recovery of upper airway reflexes shows association with<br>AEs |

ASA-PS = American Society of Anesthesiologists Physical Status;
TIVA = Total Intravenous Anesthesia; GA = General Anesthesia.
<sup>1</sup> Birth abnormality, including preterm birth and low birth weight.
<sup>2</sup> Airway sensitivity, including active or within 14 days upper respiratory infection (URI)
symptoms, asthma, living with an active smoker.
<sup>3</sup> Environmental sensitivity, including food or medication allergies, allergic rhinitis, atopic
dermatitis.

## Data quality control

This study applies the same data validation system of J-PEDIA [8] to minimize reporting and selection biases. Site-specific research leaders will be preassigned to each institutional research group before initiating data collection. This validation process is standardized for all participating sites.

The anesthetists will report all data listed in Tables 1–3 on paper-based case report forms. The research leaders at each site will validate the initial paper-based reports and check for missing information and unreported cases that should be included in this study on a daily basis. The site research leaders will collect the necessary information from the anesthesia providers assigned to the cases and/or by reviewing the anesthesia and medical records. We aim to achieve a capture rate of at least 95% for all cases at each site before initiating data collection for this study.

An operations research manual was developed by the primary and research investigators (MO and SI) to standardize the definitions of the data collected. The research operational committee will be convened regularly to confirm the definitions of the terms used in the data collection process. The site-specific research leaders will distribute the operations research manual to anesthesia providers to enhance the accuracy of the data collected at each recruited institution and minimize misclassification bias. The research collaborators can confirm uncertainties regarding data collection (such as definitions of the terms) and share information with each other using communication software (Slack®, Slack Technologies, San Francisco, California, USA). A practice period (several weeks to one month) will be scheduled for each institution to facilitate data collection by the anesthesia providers.

## Data storage and processing

The data from the paper-based data forms will be entered into the Research Electronic Data Capture system (National Centre for Child Health and Development, Tokyo, Japan) by the site-specific research leaders.

## Statistical analysis

Categorical variables will be summarized as numbers and percentages. Continuous variables will be reported as medians with interquartile ranges for non-normally distributed data. Univariate analysis will involve the use of chi-squared or Fisher’s exact test for categorical variables and the Mann–Whitney U test for non-normally distributed numerical variables. The following composite variables will be created for incorporation into the multilevel regression models: (a) respiratory comorbidity (such as respiratory support, hypoxemia, apneic events, upper airway obstruction, and laryngeal abnormalities); (b) airway sensitivity, including active or recent (within 14 days) upper respiratory infection symptoms, asthma, and living with an active smoker; (c) environmental sensitivity (such as to food or medication allergies, allergic rhinitis, and atopic dermatitis); (d) cardiovascular conditions (such as shock, cardiac arrest, congenital cardiac diseases, pulmonary hypertension); (e) physical conditions (such as American Society of Anesthesiologists Physical Status ≥ D, decreased muscle strength, preterm birth, low birth weight); and (f) gastrointestinal conditions (such as non-compliance with nil per os, full-stomach pathophysiology, nausea, or vomiting).

Multilevel multivariable regression analyses will be used to adjust for the following potential confounders as fixed effects: sex, age, body weight, American Society of Anesthesiologists Physical Status, surgical emergency, presence of full-stomach pathology, airway sensitivity, physical conditions, preoperative respiratory and cardiac comorbidities, difficult airway features, location of extubation, type of surgery, chromosomal abnormalities, use of muscle relaxant monitoring, type of anesthetics for anesthesia maintenance, intraoperative range of end-tidal CO_2_, use of muscle relaxant reversal, anesthesia provider discipline, and total number of airway-securing attempts. The variances of each institution (level 2) and type of institution (pediatric, mixed adults-pediatric university, community hospitals) (level 3) will also be adjusted. Multicollinearity will be assessed using the variance inflation factor, with a threshold of 5.

We will perform a complete case analysis. We anticipate minimal missing data because our standardized data collection system will facilitate daily checks for missing data by site-specific research leaders. The REDCap® data registration system was also established to minimize missing data. It requires the registration of most information before case registration on the REDCap® platform can be completed. Data will be analyzed using Stata V.18.0^®^ (StataCorp, Texas, USA). Two-sided *p*-values of <0.05 will be the threshold for rejecting the null hypothesis.

## Sample size estimation

Our preliminary study data showed an extubation-related respiratory AE incidence of 16%, and no laryngospasm was observed in the 112 participants. Ungern-Sternberg et al. reported perioperative respiratory AEs in 15% of the children, including laryngospasm in 4% [9]. Another randomized controlled trial reported that the incidence of laryngospasm during emergence from anesthesia in pediatric patients with a high risk of the condition who underwent adenotonsillectomy was 2% [10]. Therefore, we estimated the sample size based on an assumed lowest incidence of critical AEs (especially laryngospasm) of 2%. The estimated sample size was approximately 8500, based on a 95% Wilson CI and a margin of error of 0.3% for the incidence of AEs.

## Discussion

The prospective, registry-based, multicenter, cross-sectional study that aims to describe the real-world incidence of AEs associated with extubation and the associated risk factors in Japan.

Several large registry-based studies have been conducted in Europe and Western countries to explore the risk factors associated with airway management (such as APRICOT and PeDI studies). However, they were not specific to post-anesthetic extubation, and they did not provide detailed information on TE. Evidence from the Asian region on TE performed under general anesthesia in pediatric patients is limited.

The knowledge to develop safe TE strategies are essential to prevent critical AEs in children. Clinicians need to carefully determine the optimal anesthetic plane for TE. A previous single-center observational study showed that several awakening signs, such as facial grimace, eye opening, purposeful movement, tidal volume greater than 5 mL/kg, and conjugate gaze, were associated with a successful TE in children after emergence from a general anesthesia maintained with volatile anesthetics [6]. In this study, Templeton et al. showed the feasibility of assessment for a successful awake TE based on the classical physical findings. However, the signs for safe deep extubation and TE for TIVA maintenance for general anesthesia in children are still unknown. In addition, there is inadequate pediatric evidence regarding the patient and anesthesia risks for AEs during TE. We will reveal the signs for safe TE and the risk factors for AEs attributed to extubation based on the real-world data of clinical practice.

Some previous studies have reported difficult airway features as risk factors for reintubation [7]. However, large registry-based studies from the Asian region that have adjusted for potential confounders to determine the quantitative effect of difficult airway features on AEs during TE in children are limited. The attribution of surgery (such as intermaxillary fusion) and airway management (such as multiple airway-securing attempts) to AEs during the postoperative period of TE has not been sufficiently explored. We will investigate the potential detrimental impact of postoperative patient and airway conditions that may arise after surgical manipulations.

This study has some limitations due to its observational design. First, unmeasured confounders cannot be adjusted for. Second, selection bias may occur due to the sampling method. Reporting bias may also occur due to incorrect reports during data collection or registration. We have reviewed the literature and held repeated discussions with experienced board-certified anesthesiologists to determine the exposures, potential confounders, and institutions. To mitigate reporting bias, we have implemented a standardized system to verify the data collected by the local research leaders. This system has been used in NEAR4KIDS, which achieved a data capture rate of >95% [11]. We will use the REDCap® data registration system, which does not allow data registration to proceed if any data are missing. We have used this system in a previous J-PEDIA study and achieved successful data collection. We have conducted a pilot study to investigate feasibility and consulted research members about data collection and study methods.

Several extubation-related critical events, such as laryngospasm, are attributed to exaggerated stimulations of upper airway reflexes that are uncontrolled by the central nervous system [3,4,12].

We speculate that the period during which central nervous system inhibition resolves is the appropriate timing for successful extubation. We also believe that the abnormalities in gaze are likely related to estimating this timing. This is supported by the reported correlation between the excitatory phase and eye findings [13,14] since Guedel first proposed the concept (stage D) and its distinctive clinical findings. We consider the gaze to be a predictor of the period during which central nervous system inhibition is uncontrolled. We hypothesize that persistent cough is strongly associated with respiratory AEs. This is based on its association with enhanced expiratory reflex (cough-like reflex without preceding inspiratory flow) and its potential as a predictive sign of laryngospasm [3,12]. We consider sustained cough a predictor of severe laryngospasm and aim to classify the cough based on the associated risk.

This study can facilitate further research on the pathophysiology of airway management and upper airway reflex control system under anesthesia.

## Data Availability

The dataset will not be publicly shared due to ethical restrictions imposed by the Institutional Review Board (IRB) of the Institute of Science Tokyo, which prohibits the sharing of individual participant data with external parties. However, the detailed study protocol and statistical code are available from the corresponding author upon reasonable request.

## List of abbreviations

AEs: adverse events CO_2_ : carbon dioxide
CPAP: continous positive airway pressure
ECMO: extracorporeal membrane oxygenation
EtCO_2_: end-tidal carbon dioxide
Ex-PEDIA: Extubation in Padiatric Anesthesia
ICU: intensive care unit
PACU: post-anesthesia care unit
RAE: right angle endotracheal
TE: tracheal extubation
TI: tracheal intubation
TIVA: total intravenous anesthesia
ASA-PS: American Society of Anesthesiologists Physical Status
GA: general anaesthesia

## Declarations

### Ethics approval and consent to participate

This study will be conducted in accordance with the principles of the Declaration of Helsinki. It has been approved by the Institutional Review Board of the Institute of Science Tokyo on 7 October 2025 (Reference number: I2025-094), and local ethical approval will be obtained from each recruited institution before data collection. We have obtained adequate verbal consent from the participants for data collection.

## Consent for Publication

Not applicable.

## Authors’ contributions

MO, SI, TU, KI, FW, SK, YK, TI, and TK were involved in the study design, critical revision, and final approval of the manuscript. MO drafted the manuscript. TI and TK provided statistical expertise.

## Acknowledgements

English language editing of this manuscript was provided by Editage.

